# The NGAGE Model Developed at the Rush Alzheimer’s Disease Research Center: An Approach to Community Engagement and Inclusion in Dementia Research with Diverse Community-Dwelling Older Adults

**DOI:** 10.1101/2024.10.31.24316461

**Authors:** Crystal M. Glover, Julie A. Schneider, David A. Bennett, Lisa L. Barnes, David X. Marquez, Neelum T. Aggarwal, Sue Leurgans, Karen L. Graham, Susan Frick, Raj C. Shah

## Abstract

**Background:** One of the biggest challenges in the field of Alzheimer’s disease and related dementias (ADRD) is the severe inequitable inclusion of Black and Latino adults in clinical research studies. Despite consistent and persistent efforts, rates of participation among diverse older adults remain critically low.

**Objective:** The purpose of this paper is to set forth The NGAGE Model, one developed at the Rush Alzheimer’s Disease Research Center (Rush ADRC) to facilitate community engagement and research participation among diverse older adults.

**Methods:** The NGAGE Model consists of five steps that are conceptually distinct but overlapping in practice: 1) Networking, 2) Give first, 3) Advocate for research, 4) Give back, and 5) Evaluate. We define and describe each step. For steps 1 through 4, we calculated the number of events, number of attendees for each event, and percentages of attendees by racial and ethnic categorizations annually from July 1, 2011, through June 30, 2023, resulting in data for 12 distinct years, as provided in annual progress reports to the National Institute on Aging. For Step 5, we counted the number of persons and computed percentages of people by racial and ethnic groups who consented to our Data and Specimen Repository and enrolled in a research study.

**Results:** Over 12 years, the Rush ADRC conducted 5,362 events with 265,794 attendees. Give First activities represented the NGAGE step with both the highest number of events (n=2,247) and the most attendees (n=124,403). Among Black adults, the highest attendee percentage existed for Advocate for Research events (47%), while the highest for Latinos occurred for Give First activities (26%). Furthermore, 2,135 persons consented to the Data and Specimen Repository and 5,905 enrolled in a research study across 12 years. Higher percentages of both Black (37%) and Latino (10%) adults enrolled in research studies compared to the Repository with 21% and 7%, respectively.

**Conclusions:** The NGAGE Model facilitated community engagement and research inclusion among Black and Latino adults, particularly via Give First and Advocate for Research activities. We discuss the impacts of study milestones, staff resources, and the COVID-19 pandemic on The NGAGE Model activities and outcomes.

## Introduction

One of the biggest challenges in the field of aging is the severe inequitable inclusion of Black and Latino adults in clinical research focused on Alzheimer’s disease (AD) and related dementias (ADRD). [1–7] Insufficient representation of Black and Latino adults exists across the research spectrum, from outreach and engagement activities to participation in both observational studies and clinical trials. Simultaneously, older Black and Latino adults face a disproportionate risk of developing ADRD, with estimates ranging from 1.5 to 2 times higher risk compared to their non-Latino White counterparts.[8–12] Furthermore, ADRD devastates not only Black and Latino adults living with dementia but their entire families and communities who comprise formal and informal care networks, with dire consequences of ADRD impacting every facet of life, including current and future financial, physical, and psychological outcomes.[13–15] As such, the resounding under-representation and under-inclusion of Black and Latino adults in ADRD research severely impedes the field’s advancement. [2,16,17] More specifically, the field lacks necessary data from Black and Latino adults to understand aging experiences and ADRD trajectories in diverse communities and, thus, how to address the disparate burden of ADRD within these populations. The absence of such data severely limits the development and implementation of effective community engagement approaches and necessary informational materials regarding common topics related to aging and ADRD (e.g., managing comorbidities and phases of AD); and undermines the safety and applicability of strategies and therapeutics to diagnose, mitigate, and treat ADRD. Overall, Black and Latino adults cannot fully benefit from scientific advances and existing knowledge associated with ADRD research as findings are non-generalizable, inapplicable, or unavailable to these populations. It is paramount that a sweeping and marked shift must occur in the field regarding the inclusion of Black and Latino adults in ADRD clinical research.

Researchers have continuously put forth efforts to remedy the under-representation and under-inclusion of Black and Latino adults in ADRD research. Unfortunately, these efforts have yielded less than optimal participation levels among diverse adults.^19,21-27^ Well-documented barriers to research participation have persisted at the individual-, study-, and structural levels, including distrust of the medical system and research due to past and current mistreatment, caregiving responsibilities, cultural perceptions regarding ADRD, lack of familial support for research participation, a dearth of diverse researchers, linguistic incompatibility between study resources and communities, and inadequate transportation. [18–23]

Conversely, community engagement has facilitated, in part, existent research participation by Black and Latino adults. Researchers focused on welcoming Black and Latino adults into ADRD research have tenaciously developed community-based approaches to collaborate with diverse communities to increase research participation. [3,5,6,18,24–28]The foundation of effective community engagement consists of researchers establishing and maintaining trusting and mutually beneficial relationships with community leaders and members. These researcher-community relationships result in the bidirectional transmission of knowledge, the development and use of information and programming that is both scientifically relevant and of use to community-based audiences, current research participants as study ambassadors and collaborators, and institutional support of local community-based services and resources. [29–32] However, a recent systematic review regarding recruitment and retention with diverse older adults in ADRD research notes that related strategies vary widely by research site and largely lack a clear evaluation plan. [3] Furthermore, current recruitment and retention efforts specifically geared toward diverse older adults do not pertain to longitudinal studies focused on ADRD. [3]

The Rush Alzheimer’s Disease Research Center (Rush ADRC) has largely focused its community engagement efforts to facilitate research participation by older Black, Latino, and other minoritized populations traditionally under-included in ADRD research. Funded by the National Institutes of Health (NIH) since 1994, the Rush ADRC’s robust research portfolio mostly consists of longitudinal cohort studies focused on cognitive aging, including studies that request biospecimens and postmortem brain tissue. Initially, the Rush ADRC’s Outreach, Recruitment, and Engagement (ORE) Core provided tactical support for study recruitment and related activities. Largely through its ORE Core and support from the Patient-Centered Outcomes Research Institute (PCORI), the Rush ADRC subsequently began to foster enduring bi-directional relationships with community organizations, leaders, and members to address barriers to and facilitate increased research participation among prominent populations of older adults across the Chicago metropolitan area, including older Black and Latino adults. Hallmarks of the Rush ADRC’s approach to community engagement included community-based research activities - from recruitment events to study visits - and the creation and dissemination of evidence-based, scientific- and community-relevant, and linguistically congruent education materials.

In 2010, the Rush ADRC’s ORE Core re-examined its activities and recognized that their efforts formed the foundation of a more sustainable approach - one that is both systematic and pragmatic - to community engagement and facilitation of equitable research participation among diverse older adults. Fundamentally, the ORE Core approached community engagement by first networking with organizations that actively support diverse older adults and addressing needs identified by diverse community leaders and members then advocating for research participation. In 2011, the ORE Core formally characterized its approach to community engagement as the NGAGE Model, a community-based, participant-centered, information-focused model (See Figure 1).^31,32^ The NGAGE Model consists of five conceptually distinctive and sequential but overlapping steps: 1) Network, 2) Give first, 3) Advocate for research, 4) Give back, and 5) Evaluate. The purpose of this paper is to describe the NGAGE Model and resultant research participation in Rush ADRC and other NIH-supported studies among diverse older adults, specifically older Black and Latino adults. We developed and evaluated the process of the NGAGE Model to examine its utility as an example of effective community engagement and subsequent recruitment and retention as well as to advance recruitment science in ADRD research. As such, the NGAGE Model may inform and bolster national research efforts to facilitate representative and inclusive ADRD studies to develop and improve informational materials, treatment and risk reduction methods, and other intervention strategies and therapeutics to foster equity in aging and dementia.

**Figure 1:**
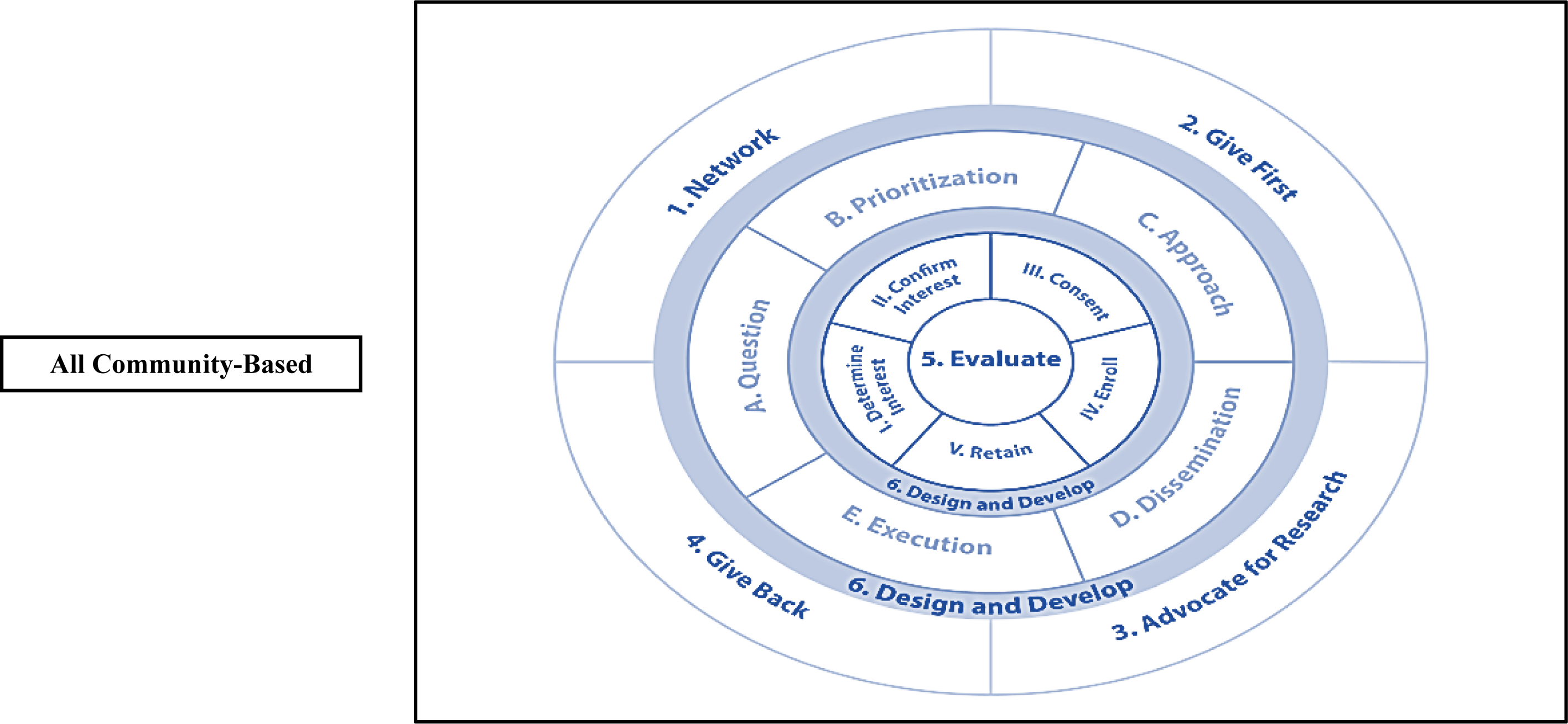
The NGAGE Model, Community and Participant Input on Study Activities, and Individual Rush ADRC Study Operation. **<u>Outer circle</u>** represents community-based events and activities associated with steps of the NGAGE Model, including Step 1: Network, Step 2: Give first, Step 3: Advocate for research, and Step 4: Give back. **<u>Middle circle</u>** represents community and participant input throughout the research process, including A. Question development, B. Prioritization of topics, C. Approach to the study, D. Dissemination of materials and study findings, and E. Execution of the study. **<u>Inner circle</u>** represents individual study activities, including the Religious Orders Study, African American Clinical Core, and Latino Core. At the core is <u>Step 5. Evaluate</u> of the NGAGE Model.

## Methods

### Participants

The Rush ADRC focuses its efforts on community-dwelling, non-demented older adults from diverse backgrounds and who are traditionally under-included in ADRD research, particularly older Black and Latino adults. The Rush ADRC’s ORE Core aims to engage with diverse communities across the Chicago metropolitan area, with its efforts guided by the NGAGE Model. Activities related to the ORE Core were approved by an Institutional Review Board (Protocol Number L91020181-AM59) at The Rush University Medical Center. Data are publicly available, de-identified, and aggregated from required annual progress reports to NIH for the Rush ADRC.

### Activities Associated with Each Step of the NGAGE Model

The first step of the NGAGE Model involves networking with leaders of community organizations and other prominent community members. Networking interactions provide insight on how to best engage with communities and “give first” (second step) to meet community-specified needs using services supported by the Rush ADRC. Community engagement leads to the third step or advocating for research. Here, ORE Core events center on providing information to community members regarding the role and value of research participation and encourage volunteering for Rush ADRC and other NIH-supported research and related initiatives. Cumulative knowledge from the Rush ADRC as well as from NIH, the Alzheimer’ Association, and other reputable scientific entities is “given back” (fourth step) to community members - regardless of consenting or not consenting to research or related participation. The Rush ADRC and its ORE Core evaluates (fifth step), in part, by examining the number of older adults who enroll in the Rush ADRC Data and Specimen Repository and those who join Rush ADRC studies and other NIH-supported initiatives. See Table 1.

**Table 1:**
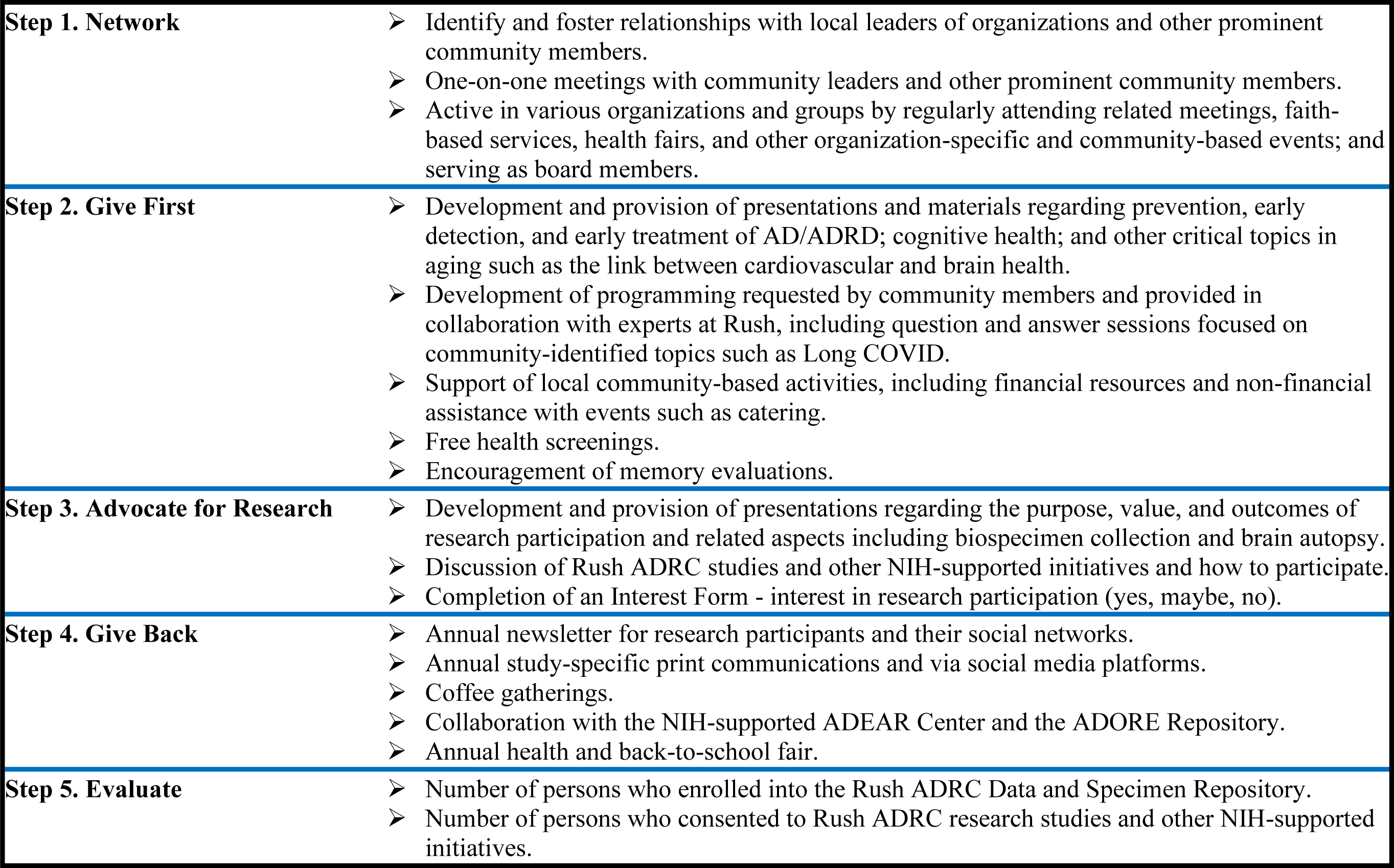
Main Events and Activities Associated with Each Step of the Network, Give first, Advocate for research, Give back, and Evaluate (NGAGE) Model Developed at the Rush Alzheimer’s Disease Research Center.

|  |  |
| --- | --- |
| **Step 1. Network** | ➤ Identify and foster relationships with local leaders of organizations and other prominent community members.<br>➤ One-on-one meetings with community leaders and other prominent community members.<br>➤ Active in various organizations and groups by regularly attending related meetings, faith-based services, health fairs, and other organization-specific and community-based events; and serving as board members. |
| **Step 2. Give First** | ➤ Development and provision of presentations and materials regarding prevention, early detection, and early treatment of AD/ADRD; cognitive health; and other critical topics in aging such as the link between cardiovascular and brain health.<br>➤ Development of programming requested by community members and provided in collaboration with experts at Rush, including question and answer sessions focused on community-identified topics such as Long COVID.<br>➤ Support of local community-based activities, including financial resources and non-financial assistance with events such as catering.<br>➤ Free health screenings.<br>➤ Encouragement of memory evaluations. |
| **Step 3. Advocate for Research** | ➤ Development and provision of presentations regarding the purpose, value, and outcomes of research participation and related aspects including biospecimen collection and brain autopsy.<br>➤ Discussion of Rush ADRC studies and other NIH-supported initiatives and how to participate.<br>➤ Completion of an Interest Form - interest in research participation (yes, maybe, no). |
| **Step 4. Give Back** | ➤ Annual newsletter for research participants and their social networks.<br>➤ Annual study-specific print communications and via social media platforms.<br>➤ Coffee gatherings.<br>➤ Collaboration with the NIH-supported ADEAR Center and the ADORE Repository.<br>➤ Annual health and back-to-school fair. |
| **Step 5. Evaluate** | ➤ Number of persons who enrolled into the Rush ADRC Data and Specimen Repository.<br>➤ Number of persons who consented to Rush ADRC research studies and other NIH-supported initiatives. |

#### Step One: Network

The first step of the NGAGE Model involves networking between Rush ADRC ORE Core staff and community leaders and prominent members located within and serving diverse communities. Staff initially identify leaders and prominent members and become consistently active in various organizations and groups within diverse communities, including regular attendance at meetings, faith-based services, health fairs, and as board members. Staff also engage in one-on-one meetings with leaders and other prominent members to further existing community-based health-related efforts. ORE Core staff learn about ongoing community efforts, and, in turn, community organizations, leaders, and members gain information about the Rush ADRC, its commitment to equity in aging, and related research efforts. At this step, the overarching goal is for the Rush ADRC ORE Core to establish relationships with local leaders and members to understand the needs and interests of diverse communities. Engagement occurs at an acceptable pace to community leaders and members, as we realize that early mistakes may jeopardize sustainable relationships.^14,41^

#### Step Two: Give First

Over time, networking can foster the development of consistent, long-term relationships between the Rush ADRC ORE Core and community leaders and members that are mutually beneficial and bi-directionally trusting. Through these relationships, the ORE Core offers “give first” activities. These events include presentations on health-related topics that are important while aging (e.g., the relationship between cardiovascular and brain health, diet and physical activity, and disaster relief and preparation) at community-based locations (e.g., park districts, libraries, and faith-based organizations); sponsorship and other resource support of community-based activities; encouragement of memory evaluation; and conducting age-friendly physical activity classes. Typically, Give First programming, including presentation topics and locations, is suggested by community leaders and members. As such, community members may also request to receive information about adjacent topics in aging, such as Long COVID and Parkinson’s disease. Here, we partner with Rush colleagues and those from nearby institutions to provide programming of interest to community members. We also pivoted Give First activities from scholarship to service during the COVID-19 pandemic, an unprecendented time, where community members requested Personal Protective Equipment (PPE) and Rush ADRC staff and other collaborators made or purchased PPE materials to provide across diverse communities. All events and activities are publicized using word-of-mouth, flyers, and social media platforms (i.e., Facebook and *X –* formally know as Twitter). Through Give First activities, the ORE Core consistently nurtures community-centered relationships, in part, by matching expressed community needs with informatinal events as well as support for community-initiated activities. Notably, Give First events may not directly tie to Rush ADRC and other NIH-supported studies.

#### Step Three: Advocate for Research

Upon establishing bidirectional and trusting relationships between the Rush ADRC and community organizations, leaders, and members - as indicated by consistent incorporation of Rush ADRC ORE Core staff in organization and group activities, we seek counsel from community leaders and members regarding the appropriate and allowable time, if any, to introduce opportunities for research participation. If permitted, the ORE Core outlines the general purpose and value of research, the need for and process of participation, anticipated outcome(s) of research and its participation, and possible implications of study findings. Then, we follow up with presentations and announcements regarding specific Rush ADRC research studies and other NIH-supported initiatives. The ORE Core aims for all presentations and correspondences regarding research opportunities to be culturally relevant and linguistically compatible with community audiences. After each presentation, attendees are asked to complete a form indicating their interest in research participation (i.e., yes, I am interested in learning more about research opportunities; maybe, I am interested in learning more about research opportunities; and no, I am not interested in learning more about research opportunities). If an attendee indicates yes or maybe, ORE Core staff contacts the person with further information. If an attendee indicates no, then we do not contact the person again at all. We encourage all attendees to share research opportunities with their social networks as well as advertise research opportunities via flyers, local radio stations, and social media platforms.

#### Step Four: Give Back

The Rush ADRC ORE Core aims to Give Back to research participants and the broader communities where they reside and represent through the dissemination of Rush ADRC research findings and related developments produced by the Rush ADRC and NIH, other NIH-funded studies, the Alzheimer’s Association, AARP, and other reputable health-related National and State entities. The ORE Core disseminates information via, in part, an annual newsletter for research participants; smaller coffee gatherings; quarterly topic-focused and annual study-specific print communications; and social media platforms. We also collaborate with the NIH-supported Alzheimer’s and related Dementias Education And Referral (ADEAR) Center and the Alzheimer’s Disease Outreach, Recruitment, and Engagement (ADORE) Repository to nationally disseminate informational and study-related materials developed at the Rush ADRC. Give Back activities also include an annual health and back-to-school fair and celebrating cultural holidays. The overall goal of Give Back activities is to support community capacity related to health and aging among diverse groups of people as well as to show appreciation for research participants and diverse communities.

#### Step Five: Evaluate

The Rush ADRC ORE Core leverages the ADRC’s Data Management and Statistical Cores to record and track the impact of NGAGE efforts for required tables in our annual NIH progress reports. We note the number and demographic characteristics of persons who: 1) consent and enter into the Rush ADRC Data and Specimen Repository from communities (i.e., not via the affiliated Memory Clinic); and 2) consent to and enroll in research studies at the Rush ADRC and other NIH-supported initiatives. We postulate that consent and enrollment into the Data and Specimen Repository and Rush ADRC and NIH-supported research studies denote linkages between NGAGE Model efforts and subsequent research participation.

### Analyses

We began systematically collecting data in 2011 regarding the NGAGE Model. For this analysis, we focus on data collected from July 1, 2011 (the formal implementation of the NGAGE model) to June 30, 2023 (the end of the fiscal year with complete data), with data broken into one-year increments (e.g., July 1, 2021-June 30, 2022), for a total of 12 years of NGAGE data. Data collected included the name of the activity or event; its corresponding NGAGE Model step (e.g., Networking); the estimated number of attendees; the percentage of attendees who belonged to the following racial categorizations: White, Black/African American, American Indian/Native American, Asian American and Pacific Islander, and multiracial; and the percentage of attendees of Latino ethnicity. Due to relatively small numbers, we collapsed the percentages for American Indian/Native American, Asian American and Pacific Islander, and multiracial categorizations into a combined racial category labeled as “Additional Groups.” For the full reporting timeframe of July 1, 2011 to June 30, 2023, we provide: 1) the number of activities and events; 2) the estimated number of attendees; 3) percentages of White, Black, and Additional Group racial categorizations of attendees; and 4) percentage of attendees of Latino ethnicity. We report these data across all NGAGE steps and for each step, cumulatively and by each reporting year.

For the Evaluate step, we aimed to understand linkages between Rush ADRC ORE Core efforts and research participation. In 2011, we began formally tracking the number and racial and ethnic identifications of individuals who: 1) consented to join the Rush ADRC Data and Specimen Repository; and 2) decided to participate in research studies at the Rush ADRC and other NIH-supported initiatives. For the Evaluate step, we report the number of persons who consented to the Data and Specimen Repository and enrolled in Rush ADRC research studies and other NIH-supported initiatives as well as racial and ethnic categorizations of participants for the full reporting timeframe and for each year.

## Results

The ORE Core supports three longitudinal cohort studies at the Rush ADRC – The Religious Orders Study (ROS), The African American Clinical Core (AA Core), and the Latino Core (LATC). ROS began in 1994 and consists of clergypersons predominately self-identified as older White adults. Since 2010, the AA Core solely includes older adults who self-identify as Black or African American. The Latino Core began in 2015 and is solely comprised of older adults who identify as Hispanic or Latino. The growth and expansion of these Rush ADRC cohort studies continue each year and require adaptations of NGAGE model activities, particularly associated with the COVID-19 pandemic and its aftermath. Furthermore, over time, the Rush ADRC provides infrastructure and resources for an increasing number of other NIH-supported initiatives and investigators. The NGAGE Model continues to inform engagement, recruitment, and retention across these diverse communities and various study needs and contexts.

### Step One: Networking

#### Number of Events

Networking comprises the first step of the NGAGE Model, where the Rush ADRC identifies and initiates contact with community-based organizations, community-serving leaders, and other prominent members of a community. Networking requires an understanding of the needs and activities of diverse communities across Chicagoland, and new and existing organizations and leaders. Across the timeframes, we observe expected fluctuations with the acceleration of Networking activities during the first three years.

Notably, one of the largest spikes in Networking occurred in Year 3, roughly two years prior to the formalization of a cohort (i.e., LATC) exclusively comprised of older Latinos in 2015. Another increase in Networking activities took place in Year 9, right before a predicted decrease due to COVID-19. See Table 2.

**Table 2.** Number of Activities and Attendees Associated with Each Step of the NGAGE Model - Network, Give First, Advocate for Research, and Give Back - From Year 1 Through Year 11*.

|  | **Year**<br>**1** | **Year**<br>**2** | **Year**<br>**3** | **Year**<br>**4** | **Year**<br>**5** | **Year**<br>**6** | **Year**<br>**7** | **Year**<br>**8** | **Year**<br>**9** | **Year**<br>**10** | **Year**<br>**11** | **Year**<br>**12** | **Total**<br>**Number** |
| --- | --- | --- | --- | --- | --- | --- | --- | --- | --- | --- | --- | --- | --- |
| **Step 1: Network** |  |  |  |  |  |  |  |  |  |  |  |  |  |
| **Activities** | 69 | 73 | 118 | 89 | 51 | 79 | 75 | 106 | 265 | 93 | 77 | 52 | 1,095 |
| **Attendees** | 2,051 | 1,706 | 7,894 | 1,826 | 1,281 | 3,228 | 1,869 | 4,608 | 7,462 | 3,020 | 1,685 | 2,264 | 36,630 |
| **Step 2: Give First** |  |  |  |  |  |  |  |  |  |  |  |  |  |
| **Activities** | 146 | 207 | 296 | 219 | 137 | 186 | 268 | 226 | 232 | 184 | 146 | 64 | 2,247 |
| **Attendees** | 20,106 | 15,819 | 21,550 | 9,869 | 9,283 | 8,923 | 12,547 | 9,175 | 9,992 | 3,625 | 3,514 | 3,529 | 124,403 |
| **Step 3: Advocate for Research** |  |  |  |  |  |  |  |  |  |  |  |  |  |
| **Activities** | 200 | 150 | 160 | 76 | 97 | 75 | 75 | 82 | 75 | 16 | 10 | 13 | 1,016 |
| **Attendees** | 11,809 | 5,812 | 6,378 | 3,471 | 3,771 | 3,257 | 4,323 | 5,466 | 5,831 | 947 | 307 | 517 | 51,372 |
| **Step 4: Give Back** |  |  |  |  |  |  |  |  |  |  |  |  |  |
| **Activities** | 84 | 109 | 112 | 65 | 53 | 43 | 44 | 59 | 87 | 81 | 68 | 68 | 805 |
| **Attendees** | 14,847 | 3,902 | 3,848 | 2,860 | 2,093 | 1,677 | 2,702 | 3,893 | 4,392 | 3,464 | 1,995 | 1,406 | 45,673 |
| **Total Number** |  |  |  |  |  |  |  |  |  |  |  |  |  |
| **Activities** | 499 | 539 | 686 | 449 | 338 | 383 | 464 | 473 | 659 | 374 | 301 | 197 | 5,362 |
| **Attendees** | 48,813 | 27,239 | 39,670 | 18,026 | 16,428 | 17,085 | 21,441 | 23,142 | 27,677 | 11,056 | 7,501 | 7,716 | 265,794 |

\* Year 1: July 1, 2011-June 30, 2012; Year 2: July 1, 2012-June 30, 2013; Year 3: July 1, 2013-June 30, 2014; Year 4: July 1, 2014-June 30, 2015; Year 5: July 1, 2015-June 30, 2016; Year 6: July 1, 2016-June 30, 2017; Year 7: July 1, 2017-June 30, 2018; Year 8: July 1, 2018-June 30, 2019; Year 9: July 1, 2019-June 30, 2020; Year 10: July 1, 2020-June 30, 2021; Year 11: July 1, 2021-June 30, 2022.

#### Number of Attendees by Racial and Ethnic Categorization

Among attendees, the number of Black adults remained consistently high during Years 1 through 3 and again in Years 10 and 11. For Years 1 through 3, Rush ADRC staff worked to maintain and grow participation in AA Core. In Years 10 and 11, Rush ADRC staff worked to identify new community-based collaborators to serve older Black adults and communities where they reside and represent in novel and necessary ways considering the COVID-19 pandemic. Conversely, a low number of Latino attendees persisted during Years 1 through 3. However, the number of Latino attendees steeply increased for Years 4 through 7, which corresponded with a heavy ORE Core focus on the assembly of LATC. Latino participation in Networking events steeply decreased during Years 8 through 11. See Table 3.

**Table 3.** Percentages of Attendees Self-Identifying as Black and Latino Adults Associated with NGAGE Model Steps - Network, Give First, Advocate for Research, and Give Back - From Year 1 Through Year 11*.

|  | Year 1 | Year 2 | Year 3 | Year 4 | Year 5 | Year 6 | Year 7 | Year 8 | Year 9 | Year 10 | Year 11 | Year 12 | Total Number |
| --- | --- | --- | --- | --- | --- | --- | --- | --- | --- | --- | --- | --- | --- |
| Attendee Race/Ethnicity**, % |  |  |  |  |  |  |  |  |  |  |  |  |  |
| **Step 1: Network** |  |  |  |  |  |  |  |  |  |  |  |  |  |
| White | 49.67 | 40.73 | 50.40 | 46.69 | 89.14 | 65.08 | 73.33 | 55.14 | 64.26 | 51.20 | 55.70 | 40.77 | 56.84 |
| Black | 34.68 | 53.13 | 41.08 | 25.32 | 9.38 | 31.24 | 23.00 | 35.51 | 31.00 | 44.54 | 37.67 | 53.00 | 34.96 |
| Additional | 15.65 | 6.14 | 8.53 | 27.99 | 1.48 | 3.68 | 3.67 | 9.34 | 4.74 | 4.26 | 6.63 | 6.23 | 8.20 |
| Latino | 6.11 | 7.05 | 7.40 | 26.17 | 74.82 | 50.92 | 46.56 | 16.38 | 38.47 | 11.91 | 7.91 | 16.38 | 25.84 |
| **Step 2: Give First** |  |  |  |  |  |  |  |  |  |  |  |  |  |
| White | 48.25 | 40.48 | 31.69 | 46.69 | 54.46 | 68.25 | 56.12 | 61.95 | 59.18 | 65.48 | 49.62 | 43.77 | 52.16 |
| Black | 37.27 | 52.88 | 66.54 | 48.52 | 38.68 | 31.04 | 40.17 | 35.18 | 36.42 | 32.58 | 48.70 | 54.82 | 43.57 |
| Additional | 14.48 | 6.64 | 1.77 | 4.78 | 6.86 | 0.17 | 2.99 | 2.87 | 4.40 | 1.93 | 1.69 | 1.51 | 4.17 |
| Latino | 19.75 | 13.70 | 13.65 | 18.96 | 34.78 | 52.64 | 32.27 | 30.81 | 43.69 | 25.71 | 17.24 | 10.08 | 26.11 |
| **Step 3: Advocate for Research** |  |  |  |  |  |  |  |  |  |  |  |  |  |
| White | 47.21 | 38.27 | 39.63 | 45.66 | 53.76 | 50.96 | 41.80 | 40.99 | 48.02 | 39.50 | 89.84 | 78.05 | 51.14 |
| Black | 48.60 | 58.66 | 59.00 | 52.60 | 44.85 | 47.93 | 56.49 | 58.16 | 48.75 | 60.48 | 5.85 | 21.95 | 46.94 |
| Additional | 4.19 | 3.07 | 1.37 | 1.74 | 1.39 | 1.11 | 1.71 | 0.85 | 3.23 | 0.02 | 4.32 | 0.00 | 1.92 |
| Latino | 16.16 | 2.64 | 12.32 | 17.71 | 27.87 | 34.11 | 12.25 | 24.23 | 35.85 | 58.54 | 29.45 | 7.16 | 23.19 |
| **Step 4: Give Back** |  |  |  |  |  |  |  |  |  |  |  |  |  |
| White | 71.37 | 63.95 | 65.51 | 61.55 | 69.41 | 79.09 | 71.73 | 67.54 | 58.43 | 58.75 | 73.58 | 74.45 | 67.95 |
| Black | 15.00 | 27.19 | 24.36 | 25.45 | 25.78 | 17.37 | 23.07 | 28.44 | 34.48 | 32.69 | 19.66 | 23.12 | 24.72 |
| Additional | 13.63 | 8.86 | 10.13 | 13.00 | 4.82 | 3.54 | 5.20 | 4.02 | 7.08 | 8.55 | 6.76 | 2.43 | 7.34 |
| Latino | 10.05 | 9.41 | 10.14 | 7.14 | 7.41 | 12.59 | 20.75 | 18.46 | 7.08 | 9.59 | 5.55 | 21.66 | 11.65 |

\* Year 1: July 1, 2011-June 30, 2012; Year 2: July 1, 2012-June 30, 2013; Year 3: July 1, 2013-June 30, 2014; Year 4: July 1, 2014-June 30, 2015; Year 5: July 1, 2015-June 30, 2016; Year 6: July 1, 2016-June 30, 2017; Year 7: July 1, 2017-June 30, 2018; Year 8: July 1, 2018-June 30, 2019; Year 9: July 1, 2019-June 30, 2020; Year 10: July 1, 2020-June 30, 2021; Year 11: July 1, 2021-June 30, 2022. \*\* Race refers to Black/African American, White, or Additional (i.e., American Indian/Native American, Asian American and Pacific Islander, and multiracial) categories; Ethnicity denotes Latino or Non-Latino.

### Step Two: Give First

#### Number of Events

Give First fosters a foundation for mutually beneficial, trusting, and lasting relationships between the Rush ADRC and various community organizations, leaders, and members. Give First activities include sponsoring an event such as financial support or food or providing an informational activity or lecture on topics of interest (e.g., heart health, physical activity, and diet). The pattern of Give Back activities during the reporting period resembles Networking events. For example, Give Back events steadily increased during the first four years of tracking, especially with preparation for recruitment into LATC. However, the number of Give First occurrences dipped, peaked once more, then decreased during the COVID-19 pandemic. See Table 2.

#### Number of Attendees by Racial and Ethnic Categorization

Attendance by Black adults remained elevated during Years 1 through 4 and peaked again in Year 11. Give First participation among Black adults follows a similar pattern as for Networking events. This suggests that Networking events were quickly followed by an ability to engage in Give First activities especially focused on Black adults. The number of Latino attendees at Give First events steadily increased from Year 1, peaking in Year 6. Subsequently, numbers for Latino attendees fluctuate and dwindle but remain robust. This pattern corresponds with efforts to establish LATC. See Table 3.

### Step Three: Advocate for Research

#### Number of Events

Once relationships are established between the Rush ADRC and community organizations, leaders, and members, researchers begin to Advocate for Research. Like Network and Give First, we tracked high numbers of Advocate for Research activities in Years 1 through 3. These increases may be related to recruiting for two established longitudinal cohort studies exclusively focused on older Black adults, including AA Core as well as the Minority Aging Research Study (MARS; PI: Dr. Lisa L. Barnes) – among communities where the Rush ADRC has long-standing relationships prior to formally tracking NGAGE efforts. Notably, Advocate for Research activities experienced a peak or higher point in Year 5, which coincided with the official start of LATC. Numbers of Advocate for Research events remained high and stable during Years 6 through 9 but steeply dropped during the COVID-19 pandemic, with numbers for Advocate for Research being the lowest of all NGAGE steps during the pandemic. See Table 2.

#### Number of Attendees by Racial and Ethnic Categorization

Black adults consistently attended Advocate for Research events, especially during Years 2 through 4 and again in Years 7 and 8. Notably, high attendance in Years 2 through 4 maps onto trends for Networking and Give First events. This may indicate that Rush ADRC staff were able to conduct Advocate for Research events sequentially and in a proximal time period as Networking and Give First activities. However, the number of Black adults who attended these events significantly dropped in Year 11. This decrease in Advocate for Research attendance among Black adults likely signals the Rush ADRC’s pivot to other NGAGE steps during the COVID-19 pandemic with less focus on showcasing and offering study participation, particularly to this population. See Table 3.

Compared to Networking and Give First events, participation in Advocate for Research events by Latino attendees began more slowly during Years 1 through 6, likely due to efforts to identify, establish, and maintain community relationships prior to explicit LATC study recruitment. Peaks for Advocate for Research events with Latino attendees existed in Years 6 and again in Year 10. Such peak years not only correspond with the beginning of LATC enrollment but also with more staff resources. See Table 3.

### Step Four: Give Back

#### Number of Events

Give Back events focus on informing research participants and communities in which they reside and represent regarding study findings and recent developments at the Rush ADRC, NIH, and other reputable scientific and health-related organizations. Research participants and related communities should not hear what is being learned from their data via news or other outlets but rather from researchers and related staff. Like previous NGAGE steps, we recorded high numbers of Give Back activities during Years 1 through 3. These increased numbers may be attributed to research findings stemming from the established AA Core and MARS cohort studies, where sustaining research participation and dissemination of findings were paramount based upon study durations. In Years 4 through 7, Give Back occurrences steadily decreased as the Rush ADRC focused more on Networking, Giving First, and Advocating for Research. However, we note an uptick in Give Back activities during the pandemic when it was, arguably, of utmost importance to raise awareness around rapid research developments as well as sustain research participation. See Table 2.

#### Number of Attendees by Racial and Ethnic Categorization

Among Black adults, overall attendance was lowest for Give Back events. It is unclear why the proportion of Black attendees was lower compared to participation in other NGAGE Model steps. Perhaps higher numbers of Black participants in Networking, Give First, and Advocate for Research events support lower numbers for Give Back activities. For Latino adults, attendance at Give Back activities remained steady across the reporting time period, with higher levels of participation in Years 6 through 8. This general pattern exists in the context of LATC simultaneously aiming to recruit and sustain participation among older Latinos; hence, concurrent peak numbers of Latino attendees in Give First, Advocate for Research, and Give Back activities. See Table 3.

### Step 5: Evaluate

For evaluating our NGAGE efforts, we obtained counts and percentages for persons who enrolled in the Rush ADRC Data and Specimen Repository and individuals who joined research studies at the Rush ADRC and those supported by NIH. Overall, more people joined research studies compared to those who enrolled in the Data and Specimen Repository, with individuals at least four times more likely to take part in research studies compared to enrolling in the Repository during Years 7 through 11. For the full reporting period, White adults comprised 58% of all participants who enrolled in Rush ADRC and other NIH-supported studies, with 37% Black adults and 10% Latinos. For Years 1 through 4, the pathway to research participation diverged for Black adults compared to Latino adults. Black adults joined research studies at higher percentages than Data and Specimen Repository enrollment. Conversely, Latino adults enrolled in the Data and Specimen Repository at higher percentages than research study participation. This pattern did reverse in Year 5, with Black adults enrolling in the Data and Specimen Repository at a higher percentage and Latinos joining research studies in higher numbers. Furthermore, Data and Specimen Repository enrollment and research participation followed a slightly different pattern in Years 8 through 10. During this timeframe, both Black and Latino adults joined research studies at higher percentages than enrolling in the Data and Specimen Repository. See Table 4.

**Table 4:** Participation in the Rush Alzheimer’s Disease Research Center (Rush ADRC) Data Repository and Rush ADRC and Other NIH-Supported Research Studies Across Participants and by Participant Race and Participant Ethnicity** From Year 1 To Year 11*.

|  | Year 1 | Year 2 | Year 3 | Year 4 | Year 5 | Year 6 | Year 7 | Year 8 | Year 9 | Year 10 | Year 11 | Year 12 | Total Number |
| --- | --- | --- | --- | --- | --- | --- | --- | --- | --- | --- | --- | --- | --- |
| **Consented and Entered into the Rush ADRC Data and Specimen Repository from the Community** |  |  |  |  |  |  |  |  |  |  |  |  |  |
| Participants, N | 232 | 357 | 384 | 418 | 241 | 150 | 116 | 44 | 26 | 45 | 59 | 63 | 2135 |
| White % | 62.50 | 63.87 | 62.50 | 71.77 | 67.22 | 80.00 | 74.14 | 86.36 | 65.38 | 75.56 | 59.32 | 71.43 | 70.00 |
| Black % | 22.41 | 13.17 | 27.34 | 22.49 | 27.39 | 10.67 | 13.79 | 9.09 | 26.92 | 11.11 | 38.98 | 25.40 | 20.73 |
| Additional % | 15.08 | 22.97 | 10.15 | 5.75 | 5.39 | 9.33 | 12.06 | 4.55 | 7.70 | 13.33 | 1.69 | 3.18 | 9.27 |
| Latino % | 14.66 | 3.92 | 5.21 | 5.98 | 5.81 | 12.67 | 16.38 | 0.00 | 3.85 | 4.44 | 8.47 | 4.76 | 7.18 |
| **Joined Rush ADRC and Other NIH-Supported Research Studies** |  |  |  |  |  |  |  |  |  |  |  |  |  |
| Participants, N | 452 | 609 | 472 | 384 | 489 | 333 | 668 | 835 | 604 | 192 | 465 | 402 | 5905 |
| White % | 70.58 | 44.83 | 54.24 | 53.39 | 71.98 | 53.45 | 55.96 | 62.28 | 42.88 | 61.46 | 65.59 | 56.97 | 57.80 |
| Black % | 28.98 | 54.68 | 45.13 | 44.79 | 19.84 | 36.94 | 37.65 | 32.34 | 52.32 | 33.33 | 26.24 | 35.82 | 37.34 |
| Additional % | 0.44 | 0.49 | 0.63 | 1.82 | 8.18 | 9.61 | 6.40 | 5.39 | 4.80 | 5.21 | 8.18 | 7.22 | 4.86 |
| Latino % | 3.54 | 1.97 | 4.45 | 5.47 | 17.38 | 23.72 | 10.03 | 10.18 | 5.79 | 14.06 | 10.97 | 10.95 | 9.88 |

\* Year 1: July 1, 2011-June 30, 2012; Year 2: July 1, 2012-June 30, 2013; Year 3: July 1, 2013-June 30, 2014; Year 4: July 1, 2014-June 30, 2015; Year 5: July 1, 2015-June 30, 2016; Year 6: July 1, 2016-June 30, 2017; Year 7: July 1, 2017-June 30, 2018; Year 8: July 1, 2018-June 30, 2019; Year 9: July 1, 2019-June 30, 2020; Year 10: July 1, 2020-June 30, 2021; Year 11: July 1, 2021-June 30, 2022.
\*\* Race refers to Black/African American, White, or Additional (i.e., American Indian/Native American, Asian American and Pacific Islander, and multiracial) categories; Ethnicity denotes Latino or Non-Latino.

## Discussion

The purpose of this study was to describe and demonstrate the utility of the NGAGE Model, an approach developed at the Rush ADRC to equitably engage, include, and sustain participation in clinical ADRD research with diverse older adults. The NGAGE Model consists of five steps that are conceptually separate but practically interconnected – Network, Give first, Advocate for research, Give back, and Evaluate. We set forth 12 years of data, from July 1, 2011 to June 30, 2023, associated with each step of the NGAGE Model. To contextualize results across all NGAGE steps, we first provide a general pattern of events and attendees across the full reporting timeframe. For the first three years, we saw a steady increase in events followed by a decrease in such activities. Notably, we restructured the Rush ADRC’s ORE Core in 2015, distributing roles throughout the Rush ADRC, followed by the addition of a new ORE co-Leader and the loss of a key staff member in 2016. We note increases in the number of events during the reporting periods between 2017 and 2019, with an all-time high across the NGAGE Model activities. We believe that the addition of new ORE and other Rush ADRC staff members and a focus on building LATC contributed to the increase in events during the 2017 through 2019 timeframes. In 2020, we experienced an expected decrease in events due to the beginning of the COVID-19 pandemic. The number of activities stabilized in 2021 forward. The maintenance of events post-2020/after the onset of the COVID-19 pandemic included a necessary and quick pivot to connecting with new organizations, leaders, and members to serve diverse older adults, including research participants.

In addition to data patterns related to NGAGE Model events and activities, we recorded numbers of attendees as well as race and ethnic categorizations of those attendees. Across NGAGE Model Steps 1 through 4, White adults comprised the majority of attendees, with slightly more than 55% being White adults, compared to Black adults (∼38%) and Latinos (∼22%). This pattern also demonstrates that at least 40% of all attendees across all events associated with Steps 1 through 4 for the total reporting period belonged to Black, Latino, and Additional racial and ethnic groups. Hence, the NGAGE Model may represent one that promotes and fosters the inclusion of diverse older adults in events and activities associated with research and related opportunities across more than a decade.

To evaluate NGAGE efforts at the Rush ADRC, we obtained counts and percentages for persons who enrolled in the Data and Specimen Repository and individuals who joined ADRC research studies and other NIH-supported initiatives. Overall, more people joined research studies compared to those who enrolled in the Repository. For the full reporting period, White adults comprised more than 50% of all research participants, with more than 30% being Black adults and 10% being Latinos. Notably, during the first 5 years of the timeframe, Black adults were more likely to join research studies while older Latinos were more likely to join the Repository. This pattern reversed in the sixth year; however, afterward, both Black and Latino adults largely joined research studies at higher percentages than enrolling in the Repository. These patterns may speak to the broader Rush ADRC context where the first half of the timeframe focused on an established AA Core, while LATC was under development with study enrollment formally occurring near the second half of the timeframe. Furthermore, in 2015, the Rush ADRC explicitly and completely de-emphasized clinic-based entry into the Repository due to predominant enrollment of non-Latino White older adults, staffing resources, and clinic priorities. The Rush ADRC simultaneously began to exclusively focus on increasing community-based engagement, recruitment, and retention to ensure research and related activities with older adults from racially and ethnically diverse backgrounds, especially older Black and Latino adults. Hence, these structural changes at the Rush ADRC likely contributed to direct enrollment in affiliated studies at the Rush ADRC, with a lower number of persons enrolling into the Repository. It is also possible that older adults are more interested in joining an active research study compared to a Repository, a possibly passive research activity. Nevertheless, the NGAGE Model was able to facilitate research participation among older Black and Latino adults during the reporting timeframe.

Previous literature has established the critical role of several factors in the decision to take part in research and related activities. [3,5,7,28,33–43] Barriers to research participation exist at the individual level, with studies being inaccessible to a person due to a lack of transportation, existent responsibilities such as caregiving, and linguistic incompatibility. These individual-level factors are inextricably tied to local and structural factors. For example, local transportation – such as insufficient or unsafe roads, sidewalks, or public transportation – is directly tied to local policy, prioritization, and investment. Academic structures, both historic and current practices, also contribute to barriers to research participation, including studies taking place at an institution of higher learning or medical center, mistreatment of Black, Latino, and other members of minoritized communities, lack of remuneration, and not sharing knowledge gained from research that may benefit communities. [3,5,10,28,40,41,44,45]Conversely, prior research has set forth two central facilitators of accessible and inclusive research – trustworthy researchers and related practices, and community-engaged research. [3,5,10,28,40,41,44,45] The field of ADRD increasingly recognizes and leverages approaches to clinical research that engage, are informed by, and based within communities traditionally inequitably included in studies. As such, it is necessary to collect and report data, especially longitudinally, associated with the reach and utility of such approaches.

We follow a long line of esteemed researchers who aimed to develop, apply, and examine community-based approaches in service of equity in ADRD clinical research. Upon this scientific foundation, the Rush ADRC conceptualized and implemented the NGAGE Model for the previous 12 years, complete with requisite data. The NGAGE Model exclusively utilized community-based and, increasingly, community-informed approaches regarding engagement, recruitment, and retention in clinical research studies, particularly focused on Black, Latino, and other communities underrepresented and underincluded in ADRD research. Data patterns indicate the ability of the NGAGE Model to guide outreach and engagement activities with diverse community organizations, leaders, and other prominent members. These relationships fostered the bidirectional trust and consistency needed to advocate for research affiliated with the Rush ADRC. Activities that focused on giving back to not only participants but also broader communities, in part, sustained research participation. Many older Black and Latino adults decided to take part in active research studies instead of signing into the data repository. While the NGAGE model represents a non-linear and iterative process, data patterns persisted through changes due to staffing resources, leadership, and the COVID-19 pandemic; thus, speaking to the durability of the NGAGE Model. Through consistent and concerted activities and events, the NGAGE Model provides a strong, pragmatic, and generalizable structure – rather than abstract or a simplified set of tactics – that may be usable and adaptable in many diverse academic contexts and communities. The NGAGE Model provides evidence-based support toward equitable engagement and inclusion with diverse older adults in clinical ADRD research. The NGAGE Model extends the field of ADRD clinical research by adding to the translational science of recruitment with diverse older adults. With this field-wide change, there will be net positive movement in required representation and inclusion in ADRD clinical research studies.

We should note that the NGAGE Model did not include the assembly and dedication of an advisory board, neither community nor participant. We decided to eschew asking community members and participants to directly assist with and focus on our research agenda but rather going to diverse communities and offering (hoping) to be a valuable service for them and supporting their decision-making regarding research. The NGAGE Model shows the possibility to facilitate diverse research participation in the absence of such advisory boards. As the use of advisory boards within ADRCs and other ADRD-focused institutions continues to grow, it is important to not lose sight of research and related efforts consistently exist within diverse communities. It remains paramount to standardize and evaluate the role, effectiveness, and experience of advisory boards from individual and community perspectives.

The current study does include limitations. First, more women took part in NGAGE model activities and events compared to men. We continue to develop NGAGE activities that may be more inclusive of men, particularly by planning to host men-only events and activities where men frequent such as barbershops. Second, while we used NIA Progress Report data for the Rush ADRC, designation of race and ethnic categories for some attendees at NGAGE Model events and activities were assessed by Rush ADRC event organizers and staff. These data were not collected from all individual attendees as this would require that people consent to enrolling into the Rush ADRC Data Repository so that we may save their Interest Form Data. Lastly, NGAGE Model events and activities took place within and surrounding the Chicago metropolitan area, with scant understanding of how this Model may work in other areas. Researchers in other geographic areas can adapt, implement, and track outcomes related to the NGAGE Model based on their research purposes. These researchers must also bolster the impact of the NGAGE Model in their work by assessing and centering the interests and needs of communities that they aim to serve through research. Cumulatively, the NGAGE Model represents a pragmatic model for community-based engagement, recruitment, and retention in service of equitable and inclusive ADRD clinical research with diverse older adults.

## Data Availability

All data produced in the present study are available upon reasonable request to the authors.

https://www.radc.rush.edu/

## Acknowledgements

We thank all community members who have attended and supported activities and events hosted by and affiliated with the Rush Alzheimer’s Disease Research Center. We thank all communities across the Chicago metropolitan area that have welcomed the Rush Alzheimer’s Disease Research Center. Last but not least, we thank all staff at the Rush Alzheimer’s Disease Center, especially those affiliated with the Outreach, Recruitment, and Engagement Core, past and present, for their dedication and tireless efforts over the years, including Traci Colvin, Israel Hernandez, Yadira Montoya, Judy Phillips, Jamie Plenge, Kyra Reynolds, and Tarisha Washington. A very special mention to Barbara Eubeler, Carol Farran, and Pamela Smith who were pivotal in developing and forming a solid foundation of ORE Core efforts.

## Author Contributions

CMG and RCS contributed to the conception and design of the research study; CMG and RCS drafted the work; CMG, DAB, LLB, DXM, NTA, SL, KLG, SW, and RCS revised the work critically for important intellectual content; CMG, DAB, LLB, DXM, NTA, SL, KLG,SW, and RCS provided approval for publication of the content; CMG and RCS agree to be accountable for all aspects of the work.

